# A prospective cohort study of prodromal Alzheimer’s disease: Prospective Imaging Study of Ageing: Genes, Brain and Behaviour (PISA)

**DOI:** 10.1101/2020.05.04.20091140

**Authors:** Michelle K Lupton, Gail A Robinson, Robert J Adam, Stephen Rose, Gerard J Byrne, Olivier Salvado, Nancy A Pachana, Osvaldo P Almeida, Kerrie McAloney, Scott D Gordon, Parnesh Raniga, Amir Fazlollahi, Ying Xia, Amelia Ceslis, Saurabh Sonkusare, Qing Zhang, Mahnoosh Kholghi, Mohan Karunanithi, Philip E Mosley, Jinglei Lv, Jessica Adsett, Natalie Garden, Jurgen Fripp, Nicholas G Martin, Christine C Guo, Michael Breakspear

## Abstract

This prospective cohort study, “Prospective Imaging Study of Ageing: Genes, Brain and Behaviour” (PISA) seeks to characterise the phenotype and natural history of healthy adult Australians at high future risk of Alzheimer’s disease (AD). In particular, we are recruiting mid-life Australians with high and low genetic risk of dementia to discover biological markers of early neuropathology, identify modifiable risk factors, and establish the very earliest phenotypic and neuronal signs of disease onset. PISA utilises genetic prediction to recruit and enrich a prospective cohort and follow them longitudinally. Online surveys and cognitive testing are used to characterise an Australia-wide sample currently totalling nearly 3,000 participants. Participants from a defined at-risk cohort and positive controls (clinical cohort of patients with mild cognitive impairment or early AD) are invited for onsite visits for lifestyle monitoring, detailed neurocognitive testing, blood sample donation, plus functional, structural and molecular neuroimaging. This paper describes recruitment of the PISA cohort, study methodology and baseline demographics.

**Author Approval:** All authors have seen and approved this manuscript.

## 1. Introduction

The neurodegenerative process underlying dementia due to Alzheimer’s disease (AD) spans several decades (Morris, 2005). Though the symptomatic burden of dementia typically occurs late in life, it is preceded by a long preclinical phase, characterized by the pernicious accumulation of neuropathology in the brain (Chiti and Dobson, 2006; Villemagne et al., 2013). This preclinical process is believed to commence decades prior to the establishment of functional decline and macroscopic brain atrophy (Dubois et al., 2016). Subtle, largely undetected changes in mood, anxiety and behaviour may accompany this process. Current therapeutic trials have usually targeted those with established AD and therefore, perhaps irreversible neurodegeneration. It is possible that earlier intervention with disease-modifying therapy may increase the chance of halting or averting neurodegenerative processes, and the ensuing burden upon the individual and society. This therapeutic window can only be identified with large-scale studies of those individuals with future risk of AD, but who currently lack substantial incipient neuropathology. The focus, therefore, of much ongoing dementia research is to elucidate early pathological changes, and to identify markers capable of predicting disease progression.

High risk cohorts for AD have typically been identified using neuropsychological tests in older adults, often recruited from “memory clinics” and with cognitive performance threshold abnormal for age (mild cognitive impairment, MCI) (Gauthier et al., 2006; Winblad et al., 2004). However, longitudinal studies of MCI cohorts are beset by several limitations. First, up to two thirds of people meeting criteria for MCI do not convert to AD within the typical time frame of a longitudinal study (Ganguli et al., 2004; Ravaglia et al., 2006). Second, those who do convert may already possess a considerable burden of AD pathology (Iturria-Medina et al., 2017). Furthermore, the cross-sectional MCI criteria fails to capture the temporal nature of cognitive decline, and lack precision because the estimated baseline performance of individuals varies greatly. Moreover, many people who do develop AD do not pass through, or present with the classic amnestic profile of MCI: Early disturbances in other domains, such as language, visuospatial and executive functions are common (Lam et al., 2013). Alternative strategies, which leverage the potential of genetics and neurobiology, are needed.

The ε4 allele of Apolipoprotein E (*APOE*) is the strongest known genetic risk factor for the common, “sporadic” (also known as late onset) AD (Saunders et al., 1993; Strittmatter et al., 1993). Large scale GWAS meta-analyses have identified at least 24 additional AD genetic risk variants (Harold et al., 2009; Hollingworth et al., 2011) (Kunkle et al., 2019; Lambert et al., 2009; Lambert et al., 2013; Naj et al., 2011). While these common genetic variants individually account for a small variance of the disease risk, their combination, as estimated by a polygenic risk score (PRS), explains a substantial amount of the heritability. A high prediction accuracy (~87% of the area under the receiver operation curve, AUC) can be achieved by a prediction model including *APOE* genotype and PRS (containing GWAS association SNPs with *p* value <0.5). The PRS adds significant predictive value over *APOE* alone (Escott-Price et al., 2015). AD PRS, with the *APOE* region excluded (PRS-no *APOE*), is a robust predictor of age of AD onset, improving the ascertainment of age of AD onset among *APOE* ε3/3 individuals by more than 10 years between the lowest and highest risk deciles (Desikan et al., 2017). Work from our own group has shown that AD PRS-no *APOE* is associated with reduced hippocampal volume in healthy older adults and those with mild cognitive impairment (MCI) (Lupton et al., 2016). The *APOE* genotype and PRS-no *APOE* both hold an association with longitudinal cognitive decline, structural MRI measures (e.g. hippocampal complex cortical thickness), and radioisotope imaging or cerebrospinal fluid identified amyloid and total tau positivity (Harrison et al., 2016; Tan, Chin Hong et al., 2017; Tan, C. H. et al., 2017).

Polygenic risk prediction therefore offers a powerful avenue to identify individuals at high risk of AD. Studying healthy mid-life adults at high genetic risk of future AD therefore offers a unique opportunity across a number of domains: to understand the neuropathological processes from their putative onset; the influence of modifiable risk factors (such as life style factors and metabolic health) on the evolution of these changes; the emergence of comorbid symptoms (such as low mood and anxiety); the variable onset of cognitive changes during conversion from high risk to AD; and the relationship between molecular, structural and functional brain changes. To do so requires ascertainment of such a cohort and detailed phenotypic characterization at regular periods, using imaging, neurocognitive assessment, genetic and biochemical characterization, lifestyle and mental health factors.

Here, we use existing cohorts with available GWAS data to establish a longitudinal cohort with elevated risk for AD to study precursors and lifestyle risk factors for AD. Our *Prospective Imaging Study of Ageing: Genes, Brain and Behaviour (PISA)* comprises the following overarching objectives:

1. Identify healthy middle-aged Australians at high risk of dementia;
2. Discover biological markers and phenotypic characteristics of early neuropathology;
3. Identify modifiable risk factors;
4. Establish a cohort of preclinical patients for future clinical trials.

## 2. Methods

### 2.1 Study Design

PISA is a prospective cohort of healthy Australians at mid-adulthood, who occupy the two tails of the genetic risk spectrum for late-onset AD. PISA participants derive from extensive in-house cohorts drawn from longitudinal studies of Australian twins and their families conducted over four decades with available genome wide genotyping data to enable a genetically enriched cohort (Figure 1). The PISA study protocol has approval from the Human Research Ethics Committees (HREC) of QIMR Berghofer Medical Research Institute and the University of Queensland.

**Figure 1.**
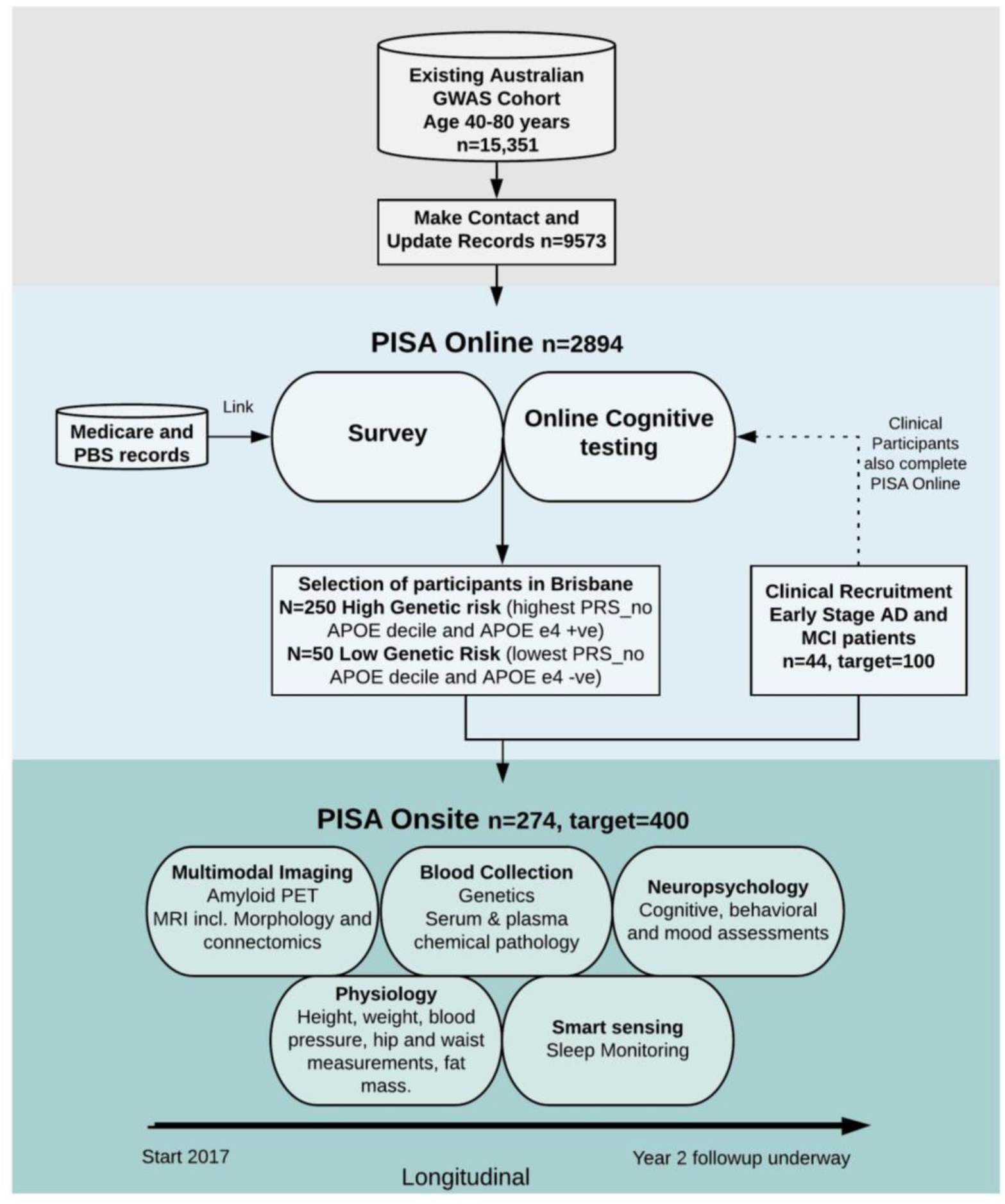
Overview of the PISA study. Sample sizes are current as of 5/12/19.

### 2.2 PISA Online

#### 2.2.1 Online survey

The population-based sample recruitment pool (N=15,351) comprises adult twins, their spouses, and first-degree relatives of twins and spouses who, over previous decades have volunteered for studies on risk factors or biomarkers for physical or psychiatric conditions and have previously been genome-wide genotyped (Benyamin et al., 2009; Heath et al., 2011). Volunteers have previously given informed consent to be contacted to take part in additional studies. These previous studies span a period of up to 40 years. Postal, telephone or email re-contact of all research participants in the age range of 40-80 years old (dob: years 1946-1986), was undertaken in order to introduce the study and update contact details. Where contact was lost, we endeavoured to obtain participant’s most recent residential address by accessing electoral roll information (available for public inspection at Australian Electoral Commission offices). Currently 9,573 (62%) participant records have been successfully updated with attempts at contact ongoing.

All participants who respond to this initial contact are invited to complete an online survey providing a global assessment of lifestyle, cognitive and behavioural function. A comprehensive online survey using Qualtrics Survey software (Qualtrics, Provo, UT, USA) allows participants to complete the survey from home (Table 1). Participants are invited by email with a link to the survey. The survey begins with an information and consent page followed by a core module that captures the central dataset (table 1 B). We also ask for permission to link participant data with MBS (Medicare Benefits Scheme) and PBS (Pharmaceutical Benefits Scheme) records. Australia has a universal health care system and this allows us to access all government subsidized medical care and prescriptions from the previous 4.5 years. Once the core module is complete, the remaining survey is made up of ten additional modules (table 1 A), which can be completed at any time in any order, with the typical time required for completion provided on the menu screen. The survey includes over 30 validated instruments (full details are listed in supplementary methods section 1). To foster collaboration, the questions were harmonized with data acquired in other national and international dementia and elderly cohorts (including the Older Australian Twin Study (OATS) (Sachdev et al., 2009), the Healthy Brain Aging (HBA) Clinic cohort eHealth survey (LaMonica et al., 2017), the Australian Imaging Biomarkers and Lifestyle (AIBL) study (Ellis et al., 2009), the European Medical Information Framework for AD (EMIF-AD) PreclinAD study (Konijnenberg et al., 2018) and the Brain Health Registry (Weiner et al., 2018).

**Table 1.** Summary of PISA online survey. A. Names of each of the 11 modules and the time taken for completion. B. Sections included in the core module Full details including instruments used and references are listed in supplementary methods section 1.

| **A. PISA Online Survey Modules** | **Average time to complete (median m:s)** |
| --- | --- |
| 1. Core Module (see B. below) | 26:00 |
| 2. Memory & Cognition | 8:31 |
| 3. Medical History | 12:27 |
| 4. Personal Wellbeing | 12:02 |
| 5. Lifestyle | 15:17 |
| 6. Personality | 15:24 |
| 7. Life Events | 15:40 |
| 8. Feelings & Emotions | 13:00 |
| 9. Physical Health | 17:04 |
| 10. Women's Health | 5:50 |
| 11. Pain | 3:37 |
| **B. PISA Online Survey Core Module Items** |  |
| Key demographics<br>Education and occupation<br>Extended family<br>Biological family medical history<br>General memory and health questions<br>Alcohol/substance abuse & smoking status<br>Patient Health Questionnaire (PHQ-9)<br>Generalised Anxiety Disorder (GAD-7)<br>Medications checklist<br>Active Australia Survey<br>Brief personal medical history (PISA exclusion criteria) |  |

#### 2.2.2 Online cognitive testing

All participants who complete the survey are invited to undertake online cognitive testing (Table 2). Participants are first invited to complete the online cognitive assessments via the Cambridge Brain Sciences (CBS) platform (Corbett et al., 2015) approximately two to three months after completion of the online survey. Participants are then approached to complete the Cogstate (Mielke et al., 2015) and Emotion Recognition (Kessels et al., 2014) tests. A second time point of online cognitive assessments is being carried out approximately one year after the baseline assessments.

**Table 2.** PISA online cognitive assessment platforms.

| Online platform | Task details | Approximate time to complete (min) | Reference |
| --- | --- | --- | --- |
| Cambridge Brain Sciences (CBS) | Twelve subtests of memory, reasoning, concentration and planning | 30 | (Corbett et al., 2015) |
| Cogstate | Online card games to test processing speed, attention (choice reaction time paradigm), visual memory (pattern separation paradigm) and working memory (n-back paradigm) | 30 (including 15 for practise session and 15 for baseline session) | (Mielke et al., 2015) |
| Emotion Recognition | Participants label emotional facial expressions presented as morphs gradually expressing one of the six basic emotions | 15 | (Kessels et al., 2014) |

#### 2.2.3 Genetic risk prediction

Genome-wide genotyping of participants within our recruitment pool was previously performed using a range of genotyping arrays including Illumina chips designed using HapMap references (317K, 370K, 610K, 660K) and more recent Illumina arrays designed using the 1KGP reference (Core+Exome, PsychArray, OmniExpress) as previously detailed (Cuellar-Partida et al., 2015; Medland et al., 2009). All datasets have been combined with strict quality control procedures and imputed to the Haplotype Reference Consortium (HRC) Release 1 reference panel (McCarthy et al., 2016).

*APOE* genotyping: The three main *APOE* alleles-ε2, ε3 and ε4 -differ at two residues (rs429358 and rs7412) and so consist of a two single-nucleotide polymorphism (SNP) haplotype. We have previously ascertained the reliability of imputed *APOE* genotypes within our in-house dataset on each different genome-wide genotyping array used, using replication genotyping of 3576 samples (Lupton et al., 2018). We used this information to assess the accuracy of the *APOE* genotype for each array used. If directly genotyped on the array or imputed with a genotype hard call imputation threshold of ≥0.9, and concordance threshold of >99.3% with our previous replication genotyping then we used the array based *APOE* genotype data. Samples not meeting these requirements were directly genotyped at the *APOE* locus using TaqMan SNP genotyping assays on an ABI Prism 7900HT and analyzed using SDS software (Applied Biosystems). SNPs were determined by allelic discrimination assays based on fluorogenic 5’ nuclease activity.

Polygenic risk score (PRS): PRS-no *APOE* was constructed from genome-wide SNP array data by summing the number of risk alleles weighted by the effect size (log odds ratio) as previously described (Lupton et al., 2017). Risk alleles were identified from the largest AD GWAS meta-analysis available at the start of the study, performed by the IGAP consortium (Lambert et al., 2013) (see Supplementary methods section 2 for details of the IGAP discovery sample). SNPs within 500kb either side of the *APOE* locus were excluded to ensure all *APOE* associated signal was removed. A large number of SNPs associated with AD risk were included in the PRS, including variants associated at a level below genome-wide significance. A threshold of *p* ≤ 0.5 was chosen for inclusion in the PRS, as this has previously been shown to have the greatest predictive value (Escott-Price et al., 2015). Linkage disequilibrium (LD)-based clumping was carried out, providing the most significantly associated SNP in each region of LD (pairwise r^2^ threshold of 0.2 and a physical distance threshold of 300kb).

### 2.3 PISA onsite

#### 2.3.1 PISA onsite overview

A subset of the population-based sample is selected for recruitment into the onsite arm of the study (PISA onsite). These include eligible individuals (criteria listed below) who are designated at high (N=250) or low risk (N=50) of AD based entirely on genetic classification. Those in the low risk group are in the lowest quintile of the AD PRS-no *APOE* (*APOE* gene region excluded) and are non *APOE* ε4 carriers. Those in the high risk group are either *APOE* ε4 carriers (including both homozygotes and heterozygotes), or those in the highest quintile of the PRS-no *APOE*. These groups, at either end of the AD genetic risk spectrum, were chosen to optimise the power to detect differences in biology and/or cognitive phenotype according to future AD risk. A detailed description of the genetic classification is given in the genetics section below.

In addition to the population-based sample, 100 early stage or preclinical AD patients are being recruited from public and private memory clinics in Australia as comparative cases. These participants are recruited to provide a cohort of early AD changes against which to benchmark findings from (high-low) risk contrasts in our healthy cohort.

The process of physical, cognitive and neurobiological ageing will be closely monitored in a longitudinal design through the use of comprehensive examinations which cover five domains of investigation: multimodal imaging (PET and MRI), genetics, neuropsychology (cognition), smart sensing (lifestyle), and physiology. In brief, participants recruited into PISA onsite undergo thorough comprehensive follow-up 2-3 years that quantify the structural and functional integrity of the brain, as well as the presence of neuropathology. Longitudinal neuropsychological, physiological and lifestyle data are also acquired at these time points. A bioinformatics database platform has been developed to capture all data, permitting multivariate and machine learning analyses to establish brain-behaviour, gene-brain, and gene-behavioural relationships and their mediation by AD genetic risk.

#### 2.3.2 Population-based cohort participant selection

Participants were approached for recruitment into PISA onsite once they had completed at least the core module on the online survey. If a participant lived within a feasible travelling distance (approximately 100km) of our testing site in at QIMR Berghofer in Brisbane, they were sent information via email or mail about the PISA onsite study. This approach has yielded a 36% response rate. Once a participant expressed interest, they were screened for metal safety and eligibility, and if they met these criteria they were invited to make an appointment. Inclusion and exclusion criteria are shown in Box 1.

###### Box 1. Criteria for recruitment of Healthy research participants into PISA onsite

Inclusion Criteria
- Recruited from existing GWAS studies at QIMR Berghofer according to their genetic risk profile (see main text). Participants previously provided a DNA sample for GWAS and gave consent to participate in other ethically approved studies.
- Aged 40-70 years
- Fluent in English
- Able to provide informed consent

Exclusion Criteria
- Any significant neurological disorder other than Alzheimer’s disease (including stroke, vascular dementia, Parkinson’s disease, Huntington’s disease, normal-pressure hydrocephalus, CNS tumour or infection, seizure disorder, multiple sclerosis, or history of significant head trauma followed by haematoma, persistent neurological deficits or known structural brain abnormalities).
- Any history of neurosurgery
- Any significant medical condition that may confound neuropsychological testing (including chronic renal failure, chronic hepatic disease, severe pulmonary disease).
- Current alcohol or substance (except tobacco) abuse or a past history of alcohol or substance (except tobacco) dependence.
- History of severe psychiatric illness or current psychiatric symptoms that may confound neuropsychological testing (including major depression with psychotic symptoms, bipolar disorder or schizophrenia).
- Females who are pregnant or breastfeeding.
- Presence of prostheses or other objects that post a risk for MRI) including pacemaker, aneurysm clips, artificial heart valves, cochlear implants, metal fragments or foreign objects in the eyes, skin or body).

#### 2.3.3 Clinical cohort participant selection

In addition to the population-based cohort, a clinical cohort is also recruited into PISA onsite. The clinical cohort is recruited via local clinicians (neurologists, geriatricians, psychiatrists and general physicians) who see patients with cognitive concerns. Recruitment sites include the following hospitals located in Brisbane: The Royal Brisbane and Women’s Hospital, The Wesley and Mater Hospitals and Neurosciences Queensland, a private practice. Referrals are made to the team of PISA clinicians who then assess the suitability of referred patients according to clinical, neuropsychological and imaging criteria. PISA aims to recruit patients in the early stages of Alzheimer’s disease or whose clinical phenotype (aMCI) is suggestive of and highly likely to predict later diagnosis of Alzheimer’s disease. To facilitate the discovery of relevant AD biomarkers, patients with significant comorbidities (including, but not limited to major psychiatric illness or cerebrovascular disease) are excluded. If the inclusion criteria (Box 2) are met, patients are discussed at a consensus meeting between PISA clinicians, before determining their suitability for enrolment. The consensus meeting involves a review of case notes, available clinician correspondence, CT, MR and metabolic imaging (including SPECT and FDG-PET but not amyloid PET), and neuropsychological test results, where available.

###### Box 2. Criteria for recruitment of Clinical research participants into PISA onsite

Inclusion Criteria
- Meet DSM-5 Criteria for Mild Neurocognitive Disorder or Major Neurocognitive Disorder of the Alzheimer’s Type or NIA-AA Criteria for Mild Cognitive Impairment **or** Dementia of the Alzheimer’s type.
- Mini-mental state examination (MMSE) >20 **and** a Clinical Dementia Rating (CDR) of 0.5 or 1.0.

*Exceptions to these criteria were made for clinically-probable cases of logopenic primary progressive aphasia (wherein language deficits might result in a lower MMSE) or posterior cortical atrophy (PCA) (wherein amnestic deficits may not be prominent)*.

- Age 40-80 years
- Fluent in English
- Able to provide informed consent

Exclusion Criteria
- Any significant neurological disorder other than Alzheimer’s disease (including stroke, vascular dementia, Parkinson’s disease, Huntington’s disease, normal-pressure hydrocephalus, CNS tumour or infection, seizure disorder, multiple sclerosis, and history of significant head trauma followed by haematoma, persistent neurological deficits or known structural brain abnormalities.
- Any history of neurosurgery
- Any significant medical condition that may confound neuropsychological testing (including chronic renal failure, chronic hepatic disease, severe pulmonary disease).
- Current alcohol or substance (except tobacco) abuse or a past history of alcohol or substance (except tobacco) dependence.
- History of severe psychiatric illness or current psychiatric symptoms that may confound neuropsychological testing (including major depression with psychotic symptoms, bipolar disorder or schizophrenia).
- Females who are pregnant or breastfeeding.
- Presence of prostheses or other objects that post a risk for MRI)including pacemaker, aneurysm clips, artificial heart valves, cochlear implants, metal fragments or foreign objects in the eyes, skin or body).

Following the first round of onsite PISA detailed neuropsychology and imaging (multi-dimensional MRI & amyloid PET) (described below), a second clinical consensus meeting is held to establish the blinded clinical diagnosis based on current criteria for MCI and AD (Albert et al., 2011; McKhann et al., 2011). Importantly, this occurs prior to divulging amyloid PET, which “unblinds” the clinicians to the presence or absence of amyloid deposition and may otherwise bias appraisal towards or away from a presumptive clinical diagnosis of AD.

#### 2.3.4 Onsite assessment overview

All participants, both population-based and clinical cohorts, attend their first visit at QIMR Berghofer in Herston, QLD. After an overnight fast, the participant arrives at an allocated time and first discusses the study in detail with a member of the research team before providing informed consent for all aspects of the study. A blood sample is donated and physical measures are collected, including height, weight, blood pressure and pulse (seated and standing), hip and waist measurements, and fat mass and percentage. Participants are provided with breakfast and are then walked the short distance to the Herston Imaging Research Facility (HIRF) where they then participate in the MRI protocol and neuropsychological testing. The timing and order of MRI and Neuropsychological assessments is organised around imaging slots and neuropsychologist availability, to maximise the number of participants that can be assessed each day. All participants are invited to return two to three months later for the MRI amyloid PET scan. There is a low drop-out rate at this stage of the study, with only 7% of participants not returning for an amyloid PET scan. When participants attend their second visit, they receive a sleep sensing device to measure resting heart rate and sleep quality over five months.

#### 2.3.5 Multimodal imaging

A summary of the MRI parameters is shown in table 3.

**Table 3.** Summary of PISA MRI sequences. *Scans acquired on the PET/MRI.

| Sequence Name | Acquisition Time(min:sec) | Matrix size (mm<sup>3</sup>) | Voxel size (mm<sup>3</sup>) | TE (msec) | TR (msec) | FA (degree) | TI (msec) |
| --- | --- | --- | --- | --- | --- | --- | --- |
| 3D MP2RAGE | 9:02 | 256×240×192 | 1×1×1 | 2.98 | 5000 | 4 / 5 | 701/2500 |
| 3D FLAIR | 7:07 | 256×256×192 | 1×1×1 | 388 | 5000 | 4 | 1800 |
| 3D 9-echo-GRE | 8:43 | 224×182×144 | 1×1×1 | 5.84<br>4.79<br>44.16 | 50 | 15 | - |
| 2D Diffusion | 10:00 | 244×244×148 | 2×2×2 | 84 | 4700 | 90 | - |
| 2D Bold-Task fMRI | 13:38 | 206×206×144 | 2.4×2.4×2.4 | 33 | 820 | 53 | - |
| 3D Dixon | 0:26 | 320x220x144 | 1.2×1.2×3.0 | 1.29<br>2.52 | 3.97 | 9 | - |
| 3D MPRAGE* | 5:03 | 256×240×192 | 1×1×1 | 2.26 | 2300 | 8 | 900 |
| rs-fMRI * | 10:05 (20:31) | 220x220x42 | 3.1x3.1x3.0 | 30 | 2680 | 90 | - |
| PC-ASL* | 9:58 | 220x220x27 | 3.4x3.4x4.0 | 13 | 5100 | 90 | - |

##### MRI (PRISMA)

Imaging data are acquired on a 3T Siemens Prisma System (Siemens Healthineers, Erlangen, Germany) with the body coil for signal transmission and a 64-channel head coil and 18-channel body coil for signal reception (software version VE11). The following sequences are acquired:

- T1-weighted structural brain images are acquired with the MP2RAGE sequence (TE/TR=2.96 *ms*/5 *s*, TI2/TI2=0.701 s/2.5 s, FA1/FA2=4°/5°, 1mm isotropic resolution, acquisition matrix 256×240×192, BW=240 Hz/Px, 3×GRAPPA acceleration) (Marques et al., 2010). MP2RAGE imaging is used to increase the consistency of the longitudinal and multi-centric data compared to that provided by the MPRAGE sequence (Okubo et al., 2016). In addition, the MP2RAGE sequence provides T1 relaxation time measurements of brain tissues that are potential biomarkers of ageing and neurodegeneration (Tang et al., 2018).
- T2-weighted fluid attenuation inversion recovery (FLAIR) images are included to assess white matter hyperintensity (WMH) burden, and parameters are as follows: TE/TR=388/5000 *ms*, FA=120°, acquisition matrix=256×256×192, 1.0 *mm* voxel isotropic.
- Multi-shell diffusion-weighted images (DWI) are acquired using parameters: TE/TR=84/4700 *ms*, FOV 244 *mm*, acquisition matrix 122 x 122, 74 slices, slice thickness 2.0 *mm*, FA=90°. The acquisition includes 12 non-diffusion weighted images (b = 0 *s/mm*^2^) as well as 20 (b = 1000 s/mm^2^), 32 (b = 2000 s/mm^2^), and 60 (b = 3000 *s/mm*^2^) unique directions. These are split across four blocks with alternating AP/PA phase encoding directions for the purpose of post-acquisition distortion correction.
- A 3D gradient-recall echo (GRE) sequence are acquired with TE1/ATE/TE9/TR=5.84/4.79/44.16/50 *ms*, FA=15°, acquisition matrix 182×224×144, GRAPPA acceleration factor=3, BW=310 Hz/Px and flow compensation only to the first echo. The GRE sequence was acquired with true-axial orientation. The phase data from individual channels were combined properly using a reference image acquired by the body coil.
- Task-based functional MRI data are acquired while participants view news clips. In brief, participants view the second half of 18 short news clips, having viewed the first half of 9 of these clips ten minutes earlier. These gradient echo data are acquired using the Siemens multi-slice acquisition with the following parameters: TR/TE=820ms/33.0ms, flip angle=53 deg, slices=72, voxel size 2.4×2.4×2.4 mm, SMS factor=6, acquisition direction PA plus two spin echo volumes with inverse PED (AP&PA) acquired for distortion correction. Brain regions activations are derived from the contrast of continuous (previous) viewing versus naïve (new) viewing. Further details of the task are provided in Supplementary Methods (section 3.1).
- In addition to the brain imaging, a two-point DIXON sequence is included to image the abdominal region between vertebral bodies T11 and L5 for assessment of visceral obesity. The acquisition is divided into two overlapping image slabs with the following parameters: TE1/TE2/TR=1.29/2.52/3.97*ms*, FA=9°, acquisition matrix 320×220×144, BW=1040 Hz/Px, voxel size 1.2×1.2 *mm* and 3.0 *mm* slice thickness.

##### PET/MRI

The positron emission tomography (PET) scans are performed on a Biograph mMR hybrid scanner (Siemens Healthineers, Erlangen, Germany) with Fluorine-18 florbetaben ([^18^F]FBB), a diagnostic radiotracer which possesses a highly selective binding for β-amyloid in neural tissue (Fodero-Tavoletti et al., 2012; Rowe et al., 2008).

With participants seated in a quiet room, 300 +/-10% MBq [18F]-Florbetaben is injected via an intravenous line inserted into a vein in the participant’s arm or hand. A 20-minute scan was acquired starting at 90 minutes post injection of [18F]-Florbetaben. The total PET/MR session takes 30 minutes to complete. PET counts are acquired using the Siemens mMR avalanche photodiodes (APDs).

For the concurrent MR, an MR UTE scan is first acquired for attenuation correction. Additionally, a T1-weighted structural image is acquired with the MPRAGE sequence (TE/TR=2.26 *ms*/2.3 *s*, T1=0.9 s, FA=8°, 1 mm isotropic resolution, matrix 256 × 240 ×192, BW=200 Hz/Px, 2x GRAPPA acceleration)(Mugler and Brookeman, 1990). Two functional MRI sequences are then acquired in parallel with PET acquisition. A subset of subjects received either a 20 minute or 10 min resting state BOLD scan is acquired with a 2D echo planar sequence (TE/TR=30ms/2680 ms, FA=90°, 3.1×3.1×3 mm resolution, matrix 220×220×42, BW= 2240Hz/Px, 223 temporal images). For subjects receiving a 10 minute rs-fMRI, this is followed by a 10 min resting state arterial spin labelling scan which is acquired with a pseudo continuous arterial spin labelling with a 2D echo planar readout (TE/TR=13ms/5100 ms, post labelling delay = 1300 ms, labelling duration 1500 ms, FA=90°, 3.4×3.4×4 mm resolution, matrix 220×220×27, 6/8 partial fourier, BW= 2298Hz/Px, 117 temporal images). These sequences allow for motion correction of the PET images and also yield resting state BOLD and CBF images for independent analyses.

All imaging data are pre-processed using a common quality controlled pipeline which integrates standard and customised steps (see Supplementary Methods sections 3.2 and 3.3 for further details)

##### Imaging data storage

DICOM images from the PRISMA and PET/MRI are collated and checked with in-house software (milxStager) for consistency with the predefined imaging protocol. DICOM data that passes consistency checks is then anonymised, converted to NIFTI and uploaded to an XNAT database (Marcus et al., 2007). RAW DICOMs are archived onto CSIRO servers. All subsequent processing is completed using data that is available on XNAT.

#### 2.3.6 Biological samples

Participants in the PISA onsite cohort have blood samples collected to a maximum of 80ml and utilised as shown in Figure 2. Serum aliquots for biochemical testing are forwarded to a clinical pathology lab (Pathology Queensland, Royal Brisbane and Women’s Hospital). The full tests carried out are described in detail in supplementary methods section 3.

**Figure 2.**
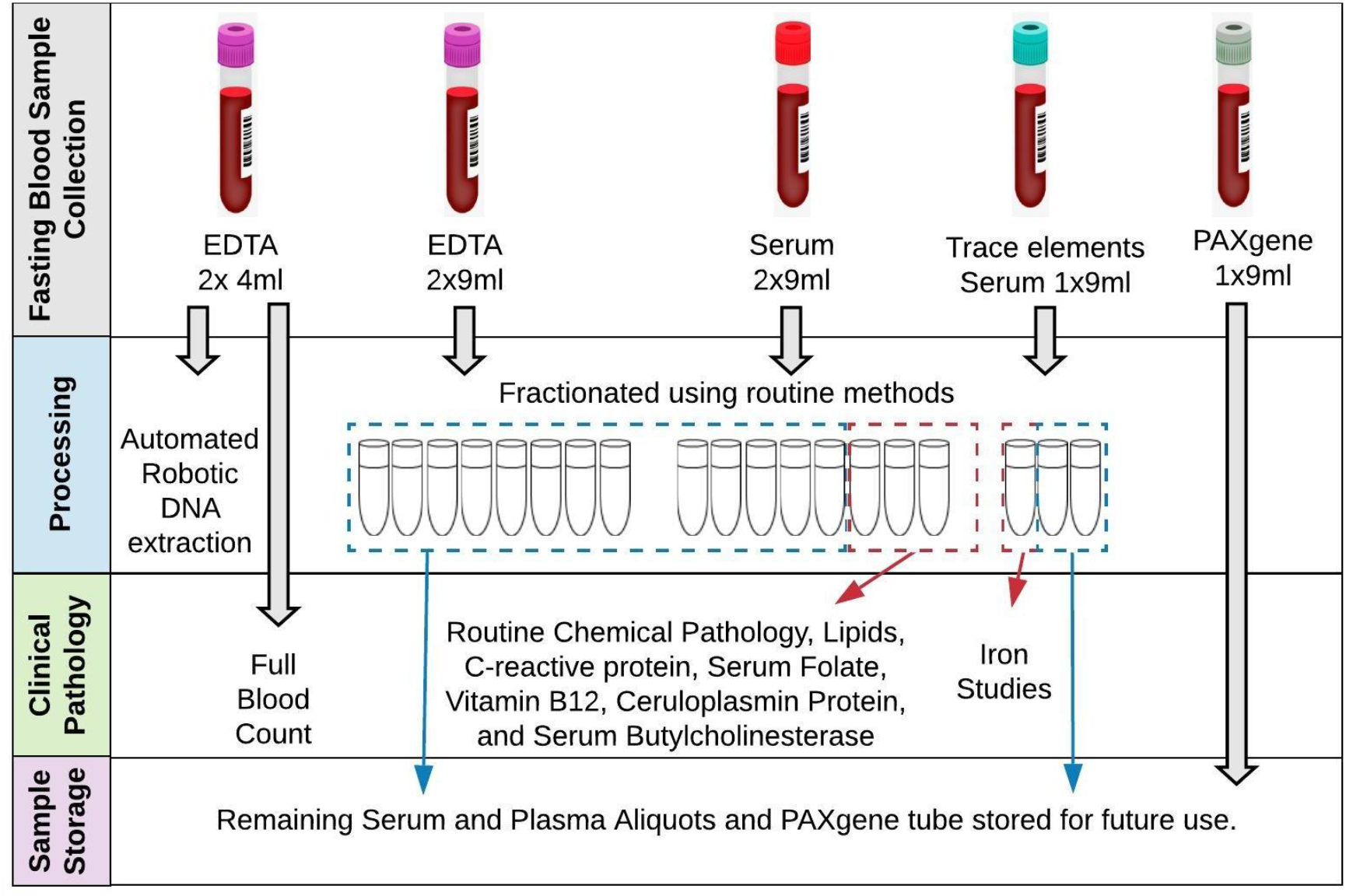
PISA Onsite blood sample collection and processing.

All samples are routinely *APOE* genotyped using the method described above, providing new data for the clinical cohort and confirming genotypes for the population based cohort. In addition clinical cohort samples are genotyped with a genome wide SNP array (Illumina Global Screening Array with Multi-disease drop-in, chip version GSAMD-24v1-0_20011747) in batches and imputed to the HRC r1.1 reference panel (McCarthy et al., 2016).

#### 2.3.7 Onsite Neuropsychology

Cognitive and mood assessments for PISA Onsite participants are conducted by trained neuropsychologists. Participants complete a range of standardized tests selected to measure the main cognitive domains that typically decline in the typical amnestic form of AD and atypical non-amnestic presentations (language, visuospatial and executive dysfunction (McKhann et al., 2011). We highlight that non-amnestic cognitive abilities are integral for memory test performance because executive functions such as attention and strategic processes are required to efficiently encode and retrieve memories and information to be recalled can be verbal or visual, implicating language and visuospatial cognitive skills. In addition, tests were chosen to enable a comparison and harmonize with other large-scale and longitudinal studies, possess robust psychometric properties, and are sensitive to longitudinal change. A select number of tests were included as proxies for cognitive reserve (e.g. reading) or that are theoretically motivated by recent neuropsychological lesion studies which implicate left/right lateral prefrontal regions in specific executive functions (e.g., Fluency and Hayling Sentence Completion Tests; (Robinson et al., 2012; Robinson et al., 2015a). Spontaneous speech samples are collected to measure language and propositional (idea) density; the number of ideas and connection or coherence between ideas (Barker et al., 2017). In the seminal Nun study, lower propositional density in younger adulthood has been shown to predict poor cognitive performance later in life and, at autopsy, confirmed AD neuropathology (Snowdon et al., 1996). All tests are administered in person, then scored and transcribed (speech samples), then summary data is entered into spreadsheets with quality control checks. Specific tests included are shown in Table 4.

**Table 4.** PISA onsite Neuropsychology assessment.

| Domain | Tests |
| --- | --- |
| General Intelligence | Wechsler Abbreviated Scale of Intelligence –2<sup>nd</sup> Edition (WASI-II). 2 sub-tests: Similarities, Matrix Reasoning) (Wechsler, 2011) |
| Memory | Episodic Verbal - Rey Auditory Verbal Learning Test (Ivnik et al., 1990) |
|  | Visual - Topographical Recognition Memory Test (Warrington, 1996) |
|  | Working Memory: Wechsler Adult Intelligence Scale – Fourth Edition (WAIS-IV; Digit Span F/B) (Wechsler, 2003) |
| Language | Naming - Graded Naming Test (Warrington, 1997) |
|  | Spontaneous speech - complex scene description – Beach Scene (Robinson et al., 2015b) |
| Literacy and Numeracy | Oral Graded-Difficulty Spelling Test (Baxter and Warrington, 1994) |
|  | Oral-Graded-Difficulty Arithmetic Test (Jackson and Warrington, 1986) |
|  | National Adult Reading Test (NART) (Nelson and Willison, 1991) |
| Executive Functions | Word fluency- FAS; Animals (Tombaugh et al., 1999) |
|  | Stroop Test (Victoria version) (Troyer et al., 2006) |
|  | Hayling Sentence Completion Test (Burgess and Shallice, 1997) |
| Psychomotor Speed | Symbol Digit Modality Test (Smith, 1973) |
| Visual Perception | Cube Analysis (VOSP) (Warrington and James, 1991) |
| Attention | TEA: Telephone Search; Dual Task (Robertson et al., 1994) |
| Social Cognition and Emotion Recognition | Mini-SEA emotion evaluation (Bertoux et al., 2014) |
|  | TASIT-S part B Sarcasm (McDonald et al., 2017) |
| Current Mood Symptoms | Hospital Anxiety and Depression Scale (HADS) (Snaith and Zigmond, 1994) |

All participants are asked screening questions about everyday function in activities and changes to functional or cognitive capacity to corroborate that they are in the healthy or clinical cohort. The length of assessment is typically two hours for healthy participants and three hours for the clinical cohort.

#### 2.3.8 Smart Sensing

Recent research demonstrates a bidirectional relationship between sleep and AD pathology, with supporting evidence showing that changes in sleep patterns occur from the preclinical stage of AD (Ning and Jorfi, 2019). The co-occurrence of sleep disturbances and amyloid-beta (Aβ) accumulation suggest the importance of monitoring sleep patterns in adults who are at high-risk of developing neuro-degenerative disorders later in life.

Polysomnography (PSG) is the gold standard in measuring sleep parameters. However, there are several limitations in using PSG for longitudinal sleep monitoring. It is expensive, intrusive, and may lead to disturbed sleep patterns, due to an unfamiliar laboratory environment. Pervasive or ubiquitous sensors are an alternative choice for measuring sleep on a long-term basis due to low cost, and minimal obtrusion in a participant’s usual environment.

PISA onsite participants are given a contact-free sleep monitoring device when they return on-site for their PET scan. The EMFIT QS (Quantified Sleep) (Emfit, Filand) (Ranta et al., 2019) sleep monitor consists of a thin ferroelectret sensor that is placed underneath a bed mattress to perform at-home monitoring of sleep patterns. During their visit, participants are provided with verbal and written instructions on how to set up and use the device. The EMFIT QS connects to the participants’ home Wi-Fi and regularly transmits data to the research team. The device collects continuous resting heart rate and respiration rate, sleep stage estimates, sleep duration, sleep latency, waking rate and number of bed exits throughout the night. The heart rate and respiration rate measured by EMFIT QS has been validated against PSG measurements (Ranta et al., 2019).

So far, 137 PISA participants have been offered an EMFIT QS device. Of these, 68 participants completed the study, 38 participants declined due to Wi-Fi availability and/or privacy concerns, and 31 participants are currently using the device. The participants are given a reply-paid postage bag with the device and instructed to return the device when 5 months of data collection has been completed.

The aim of the smart sensing stream is to understand the changes in sleep pattern across the spectrum of AD, from the preclinical stage to late-onset AD, and to investigate the impact of altered sleep in disease progression in AD. To do so, we introduce a sleep disruption index as a quantitative measure for significant changes in sleep patterns based on the collected data from the EMFIT QS.

#### 2.3.9 Bioinformatics

PISA phenotypic data is collated into a central RedCAP database (Harris et al., 2009). This includes basic MRI-derived measurements (such as hippocampal volume, white matter lesion volume, and ventricular volume) from imaging pipelines which are uploaded into RedCAP either as batch uploads or automatically at the end of the processing pipeline. Any manual annotations relevant to imaging acquisition (e.g. subject was claustrophobic, scan was cancelled after 20mins) are also uploaded. Results of the blood tests are automatically uploaded from HL7v2 files received from the pathology lab, preventing the need for manual entry and reducing data entry errors. All Additional PISA onsite phenotypic data are batch uploaded into RedCAP. Measurement data in RedCAP is periodically locked down and cross checked to ensure no errors have propagated during measurements and data entry. This is carried out both manually and via automated scripts which examine the distribution of values and mark any outliers to be cross checked. Data is made available for download to PISA investigators and collaborators using a web-based portal.

## 3. Results

### 3.1.1 Results overview

We give an overview of our current sample sizes and demographic data as of the 5^th^ December 2019 from participants recruited for PISA online and the baseline of PISA onsite. Recruitment for both is ongoing, and the follow-up time point for PISA onsite (two years after the baseline visit) is now underway.

### 3.1.2 Results PISA Online

The online survey was launched on the 4^th^ of July 2017. The current participation rate is 30%. To date, 2,894 participants have taken part in the core survey (2,789 to completion) with 76% of those going on to complete all 11 modules (Figure 3). The demographic information for participants who have completed the core module is given in Table 2. Ethnicity is almost completely Caucasian due to a prerequisite for Caucasian ancestry in the previous genetic studies which form the recruitment pool. Sample size will increase as the follow up of non-responders is ongoing, and includes provision of paper copies of the questionnaire if required.

**Figure 3.**
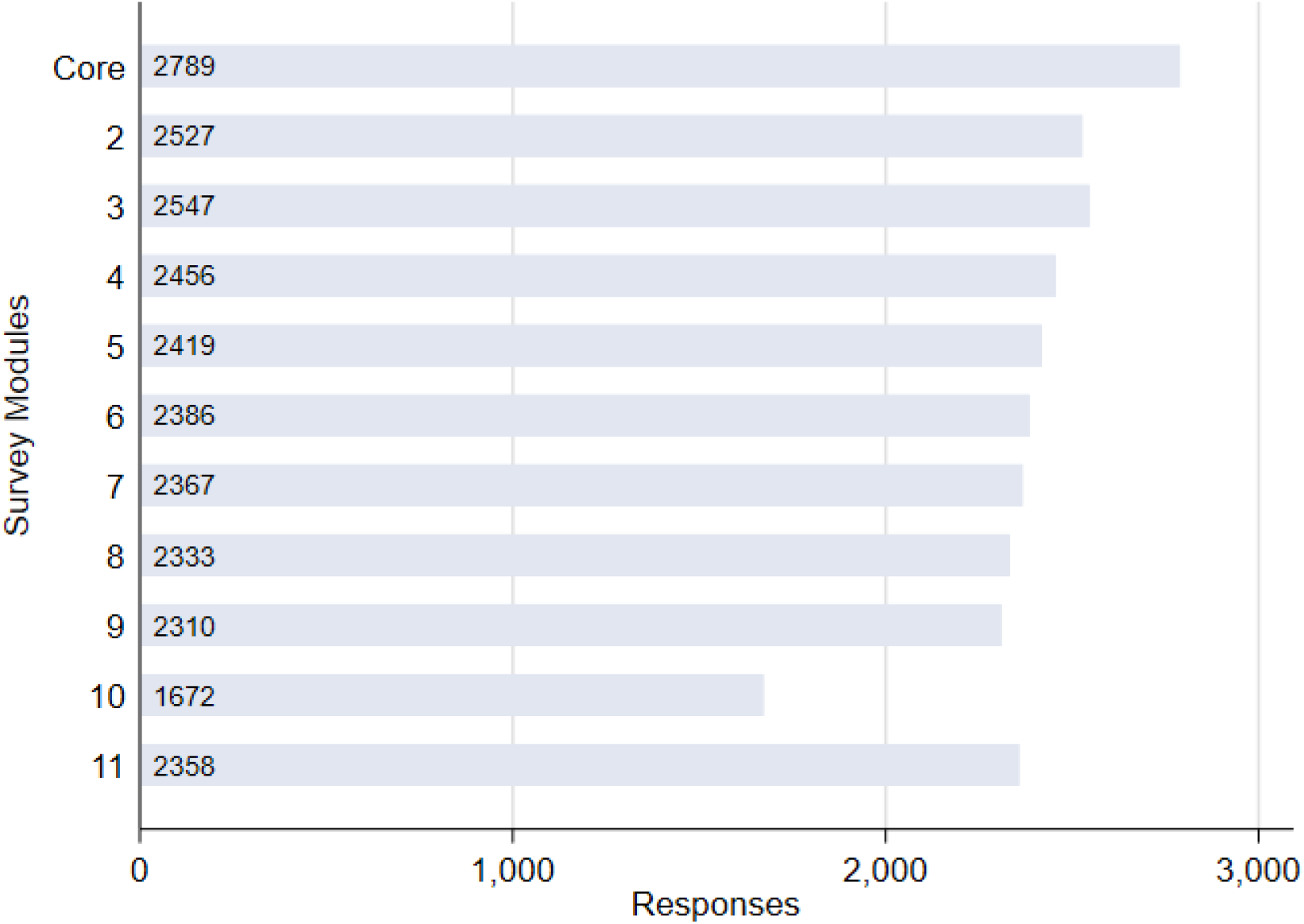
Participation in PISA online survey by module as of 05/12/19. (see table 1 for module information and supplementary table 1 for question details. Module 10 collects information on women’s health and therefore shows reduced participation because is completed by women only).

Participants who have completed the online survey are also approached to complete online cognitive assessments. Currently 1658 participants have taken part in the CBS assessment (63% participation rate from 2,647 who have been approached), 347 in Cogstate (26% of 1354 approached), and 630 completed the Emotion recognition task (46% of 1354 approached), with recruitment and follow-up ongoing.

### 3.1.3 Results PISA onsite baseline

To date 274 participants have completed baseline onsite assessments for PISA onsite. Of these 44 are clinical participants and 223 are population based (selected from those who have completed the online survey). Participant demographics are shown in Table 5.

**Table 5.**
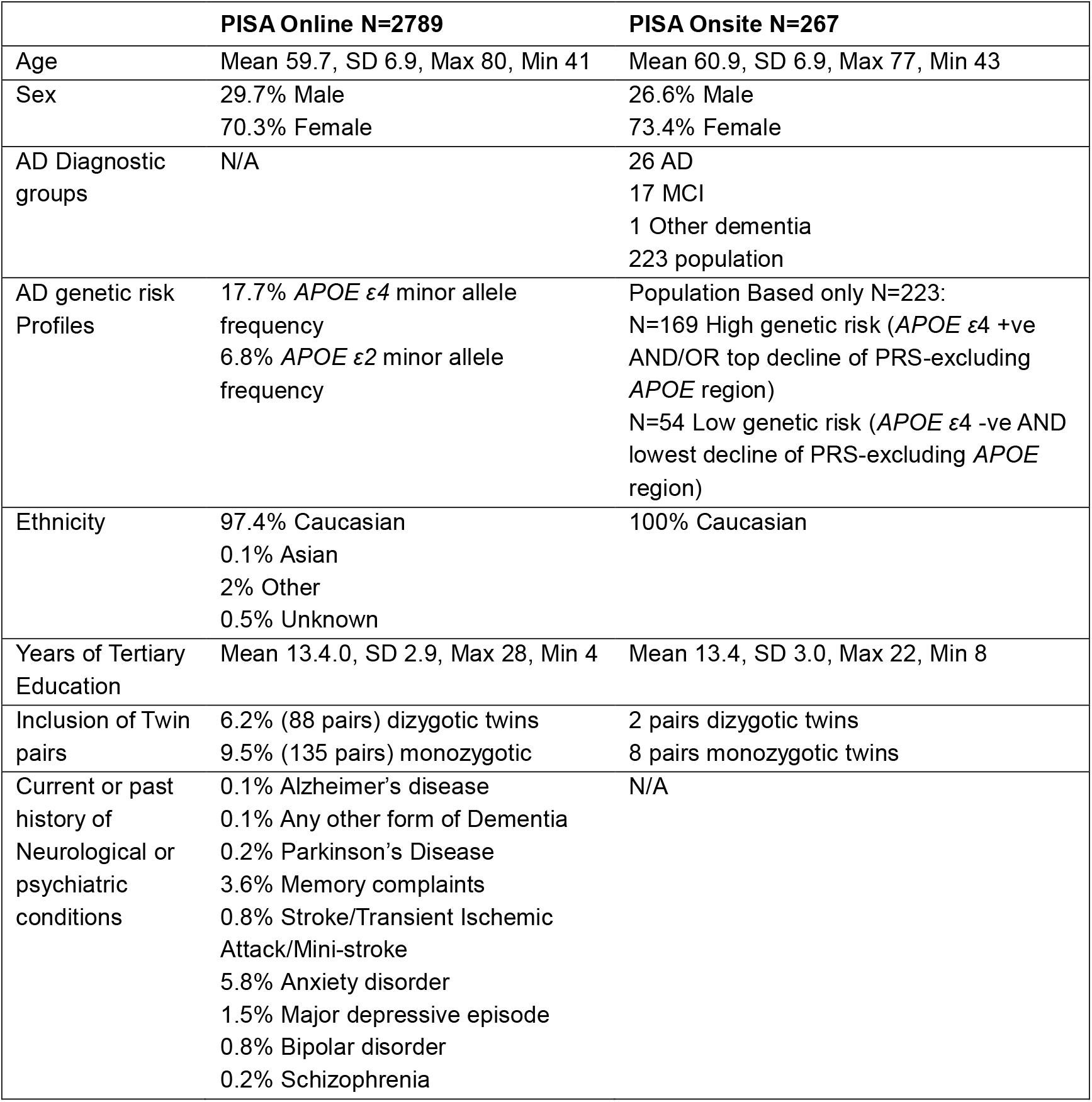
Participant demographics for PISA as of 05/12/19. For PISA online data shown for all individuals who have completed the core module of the online survey.

|  | **PISA Online N=2789** | **PISA Onsite N=267** |
| --- | --- | --- |
| Age | Mean 59.7, SD 6.9, Max 80, Min 41 | Mean 60.9, SD 6.9, Max 77, Min 43 |
| Sex | 29.7% Male<br>70.3% Female | 26.6% Male<br>73.4% Female |
| AD Diagnostic groups | N/A | 26 AD<br>17 MCI<br>1 Other dementia<br>223 population |
| AD genetic risk Profiles | 17.7% *APOE* ε4 minor allele frequency<br>6.8% *APOE* ε2 minor allele frequency | Population Based only N=223:<br>N=169 High genetic risk (*APOE* ε4 +ve AND/OR top decline of PRS-excluding *APOE* region)<br>N=54 Low genetic risk (*APOE* ε4 -ve AND lowest decline of PRS-excluding *APOE* region) |
| Ethnicity | 97.4% Caucasian<br>0.1% Asian<br>2% Other<br>0.5% Unknown | 100% Caucasian |
| Years of Tertiary Education | Mean 13.4.0, SD 2.9, Max 28, Min 4 | Mean 13.4, SD 3.0, Max 22, Min 8 |
| Inclusion of Twin pairs | 6.2% (88 pairs) dizygotic twins<br>9.5% (135 pairs) monozygotic | 2 pairs dizygotic twins<br>8 pairs monozygotic twins |
| Current or past history of Neurological or psychiatric conditions | 0.1% Alzheimer's disease<br>0.1% Any other form of Dementia<br>0.2% Parkinson's Disease<br>3.6% Memory complaints<br>0.8% Stroke/Transient Ischemic Attack/Mini-stroke<br>5.8% Anxiety disorder<br>1.5% Major depressive episode<br>0.8% Bipolar disorder<br>0.2% Schizophrenia | N/A |

## 4. Conclusion and limitations

The primary outcome of the PISA study is to establish a longitudinal cohort of healthy mid-life individuals at high genetic risk of dementia. This will be a unique international resource – only enabled by the recent breakthroughs in genetic analysis – and will yield new possibilities for basic and translational research. This protocol and cohort are of broad significance for other investigations – particularly clinical trials in dementia prevention.

Using advanced structural, functional, and molecular imaging technologies, we will elucidate the neurobiological features associated with high risk for dementia and, in particular, identify those changes associated with the initial onset of cognitive impairment in those high-risk participants as they begin the transition to AD. Together with genetic risk prediction, such knowledge has clear potential to develop prognostic markers for dementia development.

While we study the interplay between genetic and environmental factors for dementia, we also aim to identify those risk factors (e.g. lifestyle) that could be modified through intervention. The multimodal imaging, behavioural, neuropsychological and sleep data will be a unique resource to assist in understanding the complex and dynamic relationship between neurobiological antecedents of dementia at high risk and the subsequent progression to cognitive impairment.

Limitations of the PISA study include that the ‘population based’ cohort is selected from a database of previous research participants who originally consented to be part of a prior research study, allowed re-contact and then consented to take part in PISA. This multi-stage recruitment process will likely inflate any selection bias related to willingness to participate in research studies. In addition, characteristics of the original studies’ participant selection will be reflected in PISA participants including geographical locations, the inclusion of twins and relatives of twins, the large proportion of female participants (due to increased rates of females volunteering for research over males) and the predominance of Australians of Caucasian ethnicity. For the PISA onsite population cohort individuals who are at both high and low genetic risk of AD were purposely selected. Therefore, rather than a traditional case/control design where the controls are representative of the normal population, this method reflects a kind of ‘extreme sampling’ in terms of genetic risk.

## Data Availability

Following completion of each wave (baseline, follow-up) and appropriate quality control, de-identified PISA data will be made available to other research groups upon appropriate request and evidence on institutional ethics approval for local data storage, analysis and security.

## Acknowledgments

PISA is funded by a National Health and Medical Research Council (NHMRC) Boosting Dementia Research Initiative - Team Grant [APP1095227], to MB, NM, GB, SR, CCG, OS, OA, GAR, NP. MKL is supported by a Boosting Dementia Leadership Fellowship [APP1140441]. GAR is supported by a Boosting Dementia Research Leadership Fellowship [APP1135769].

The population based sample for PISA comprises of research participants who have taken part in genetic epidemiology studies led by NM (with colleagues and collaborators) at QIMR Berghofer and elsewhere over the past 40 years. Demographic and contact information as well as genome-wide SNP chip data were utilised from these previous studies for participant selection and recruitment. Principal sources of funding for these early studies were from grants to NGM from Australian NHMRC and to Andrew Heath and Pam Madden (Washington University, St Louis) from NIH (mainly NIAAA and NIDA) and we gratefully acknowledge these contributions.

We thank the International Genomics of Alzheimer’s Project (IGAP) for providing AD meta-analysis summary results data for these analyses. The investigators within IGAP contributed to the design and implementation of IGAP and/or provided data but did not participate in analysis or writing of this report. IGAP was made possible by the generous participation of the control subjects, the patients, and their families. The i–Select chips were funded by the French National Foundation on Alzheimer’s disease and related disorders. EADI was supported by the LABEX (laboratory of excellence program investment for the future) DISTALZ grant, Inserm, Institut Pasteur de Lille, Université de Lille 2 and the Lille University Hospital. GERAD was supported by the Medical Research Council (Grant n° 503480), Alzheimer’s Research UK (Grant n° 503176), the Wellcome Trust (Grant n° 082604/2/07/Z) and German Federal Ministry of Education and Research (BMBF): Competence Network Dementia (CND) grant n° 01GI0102, 01GI0711, 01GI0420. CHARGE was partly supported by the NIH/NIA grant R01 AG033193 and the NIA AG081220 and AGES contract N01–AG–12100, the NHLBI grant R01 HL105756, the Icelandic Heart Association, and the Erasmus Medical Centre and Erasmus University. ADGC was supported by the NIH/NIA grants: U01 AG032984, U24 AG021886, U01 AG016976, and the Alzheimer’s Association grant ADGC–10–196728.

